# Extended effect of short-course azithromycin for the treatment of diarrhoea in children on antimicrobial resistance in nasopharyngeal and intestinal bacteria: Study Protocol for the antimicrobial resistance sub-study of the multicountry AntiBiotics for Children with Diarrhea (ABCD) trial

**DOI:** 10.1101/2020.07.17.20156224

**Authors:** The ABCD Study Team, T Alam, D Ahmed, T Ahmed, MJ Chisti, MW Rahman, A Chauhan, S Deb, P Dhingra, U Dhingra, A Dutta, A Keshari, A Pandey, S Sazawal, S Belanger, K Kariuki, S Karuiki, P Pavlinac, B Singa, Judd L Walson, N Bar Zeev, J Cornick, Q Dube, B Freyne, V Maiden, C Ndamala, L Ndeketa, R Wachepa, H Badji, JP Booth, F Coulibaly, F Diallo, F Haidara, K Kotloff, D Malle, A Mehta, S Sow, M Tapia, S Tennant, R Anjum, A Hotwani, A Hussain, P Hussain, F Kabir, Farah N Qamar, S Shakoor, T Yousafzai, C Duggan, U Kibwana, R Kisenge, C Lomward, K Manji, S Somji, C Sudfeld, P Ashorn, R Bahl, Ayesha De Costa, J Simon

## Abstract

**Introduction:** Antimicrobial resistance (AMR) is a major public health challenge worldwide, threatening the important gains that have been made in reducing mortality due to infectious diseases. Despite current World Health Organization guidelines restricting antibiotics to a small subset of children with dysentery or suspected cholera, many children with diarrhea continue to be treated with antibiotics. We aim to determine the impact of a 3-day course of azithromycin on the risk of AMR at 90 and 180 days after treatment, among a subset of children and their household contacts enrolled into a multi-country, randomized, double-blind, placebo-controlled clinical trial of azithromycin children under 2 years with diarrhea in low income settings,

**Methods and analysis:** The AntiBiotics for Children with Diarrhea (ABCD) trial is testing the efficacy of a 3-day course of azithromycin, compared to placebo, in reducing mortality and linear growth faltering in the subsequent 6 months among 11,500 children aged 2-23 months of age across multiple sites in Bangladesh, India, Kenya Malawi, Mali, Pakistan and Tanzania with diarrhea and one or more of the following; dehydration, severe stunting, or moderate wasting (https://clinicaltrials.gov/ct2/show/NCT03130114). A sub-set of enrolled children are randomly selected to participate in a sub-study of AMR. A fecal sample (stool or rectal swab) will be collected at baseline from all enrolled children. A fecal sample and a nasopharyngeal (NP) swab will be collected at day 90 and 180 after enrolment from participating children and a close household child contact. *Escherichia coli* and *Streptococcus pneumoniae* will be isolated and Minimum Inhibitory Concentration for azithromycin and other commonly used antibiotics will be determined and compared between trial arms.

**Ethics and dissemination:** This study was reviewed by an independent ethical review committee. Dissemination of results is planned to local and international policy makers and the public.

**Registration details (Parent ABCD trial):** https://clinicaltrials.gov/ct2/show/NCT03130114

**Strengths and limitations of this study (3-5 points):**

✤ This study will provide evidence from a randomized controlled trial regarding the risk of short term azithromycin use on resistance to azithromycin and selected commonly used antibiotics, 90 and 180 days after administration in treated children and their household contacts. Few RCTs of antibiotics for diarrhoea have provided such long-term follow-up and close contact data, both of which are key to understanding the potential risk of short-term antibiotic use in the context of diarrhoea.
✤ This study will also provide data on antibiotic resistance from multiple countries in sub-Saharan Africa and Asia where availability of such data is limited.
✤ *Escherichia coli* and *Streptococcus pneumoniae* will be used as indicator organisms to monitor the impact of empiric antibiotic azithromycin administration on the development of resistance in bacteria colonising the gut and nasopharynx respectively – both are suitable for this purpose as they have pathogenic potential and are also commensal organisms which may act as reservoirs of transmissible genetic resistance elements.
✤ With only two follow-up visits at 90 and 180 days, lack of culturing of other bacterial pathogens, and minimal collection of information on other antibiotic use during follow-up, this study will not evaluate impact of azithromycin beyond 180 days, the impact on other pathogenic bacteria, nor the added impact of the use of other antibiotics on resistance profiles

## Introduction

Diarrheal disease kills over 500,000 children under 5 years annually, with the majority of these deaths occurring in sub-Saharan Africa and South East Asia.^1^ Diarrhea, and the enteric infections that cause diarrhea, also contribute to linear growth faltering among survivors.^2-4^ The current World Health Organization (WHO) recommended management guidelines for acute diarrhoea (rehydration, supplemental zinc, feeding advice and appropriate follow-up) ^5^ have contributed to significant reductions in diarrhoea-associated mortality^6^. These guidelines do not suggest a role for antibiotics except in case of bloody diarrhoea (as a proxy for shigella or other invasive bacterial infections) or suspected cholera.

However, approximately 40-80% of all children with self-limiting diarrhea currently receive antibiotics, despite WHO guidelines recommending antibiotics only to a small subset with dysentery or cholera.^7,8^ Antibiotics are widely accessible in most communities in low resource settings through pharmacies and informal vendors. In clinics and hospitals, scarce diagnostic resources and consequent therapy based on clinical syndromes that are nonspecific for serious bacterial infections (i.e. therefore likely to capture viral, parasitic, and self-limiting illnesses) also drive antibiotic consumption, which is a key factor in the promotion of anti-microbial resistance (AMR).^9^ Importantly, antibiotics are increasingly used in animal husbandry^10,11^ contributing further to the spread of resistance.

Overuse of antibiotics is associated with increased rates of antibiotic-resistant bacteria, unnecessary costs, and significant incidence of adverse events. The growing burden of antibiotic resistance threatens the substantial progress that has been made. Moreover, the spread of *Enterobacteriaceae* that produce extended-spectrum β-lactamases (ESBLs) and other multidrug-resistant (MDR) organisms in both community-based and hospital-based populations threatens the efficacy of many first line antibiotics that are used to treat diseases other than diarrhea, such as bacteremia, meningitis, and pneumonia.^12,13^

The Antibiotics for Children with Diarrhea (ABCD) trial is a double-blind, individually randomized placebo-controlled trial testing whether empiric azithromycin, administered to children at high-risk of death, reduces risk of death and linear growth faltering in the months following the diarrheal episode. This trial is based on evidence that enteric bacteria, after rotavirus, are the important causes of diarrhea-attributable deaths^14-17^ and that many of these bacteria are associated with linear growth faltering.^2,18^ Data from trials for acute respiratory illness provided clear evidence for when antibiotics were of benefit in the management of high risk children. These data and the resultant guidelines led to decreases in indiscriminate antibiotic use and reductions in resistance among isolated pathogens^19,20^.

Recent cluster trial evidence suggests a mortality benefit associated with single-dose azithromycin in high-mortality regions of sub-Saharan Africa.^21,22^ Any clinical benefit of azithromycin needs to be weighed against the potential increase in antibiotic resistance associated with an antimicrobial intervention. We seek to determine whether children treated with azithromycin for diarrhea have a higher prevalence of antibiotic resistance in gut (*Escherichia coli [E*.*coli]*) and nasopharyngeal (*Streptococcus pneumoniae [S. pneumoniae]*) bacteria compared to children who will receive placebo 90 and 180 days following study drug administration. We will also evaluate resistance in the household contacts of enrolled index children at the same timepoints.

## Methods and analysis

### Aim

The primary objectives of this nested sub-study of the ABCD trial are:

1. To determine whether the prevalence of azithromycin resistance in strains of *E. coli* isolated from stools, and *S. pneumoniae*, isolated from nasopharyngeal swabs, is no different between children treated with 3-days of azithromycin or placebo in a randomly selected sub-sample of children enrolled in the ABCD trial, at 90 and 180 days after treatment. Resistance to other antibiotic classes will be described.
2. To determine whether the prevalence of azithromycin resistance in strains of *E. coli* and *S. pneumoniae* isolated from the siblings or close household contacts (children under five years of age living in the same household under the care of the same primary caregiver) of the sub-sample of participating children in the objective above, is no different between contacts of children treated with 3-days of azithromycin or placebo, at 90 and 180 days after treatment. Resistance to other antibiotic classes will be described.

### Parent Trial

#### Design

A random subset of 15% (n=1750) of children enrolled in the ABCD trial ^23^ (n=11500) will be assigned to participate in this nested sub-study (fig 1). Random assignment into the AMR sub-study will be determined by the World Health Organization (WHO) Central Coordinating Office as part of the intervention randomization code, stratified by site and intervention arm. Children enrolled in the AMR sub-study will have a stool sample (or rectal swab) collected and processed at baseline, and a fecal sample and a NP swab collected and processed for culture and resistance testing at day 90 and at day 180 after enrolment. In addition, a stool sample (or rectal swab) and a NP swab will also be collected from a sibling or other close household contact (child contact) of the enrolled child (index-child) at day 90 and day 180.

**Figure 1:**
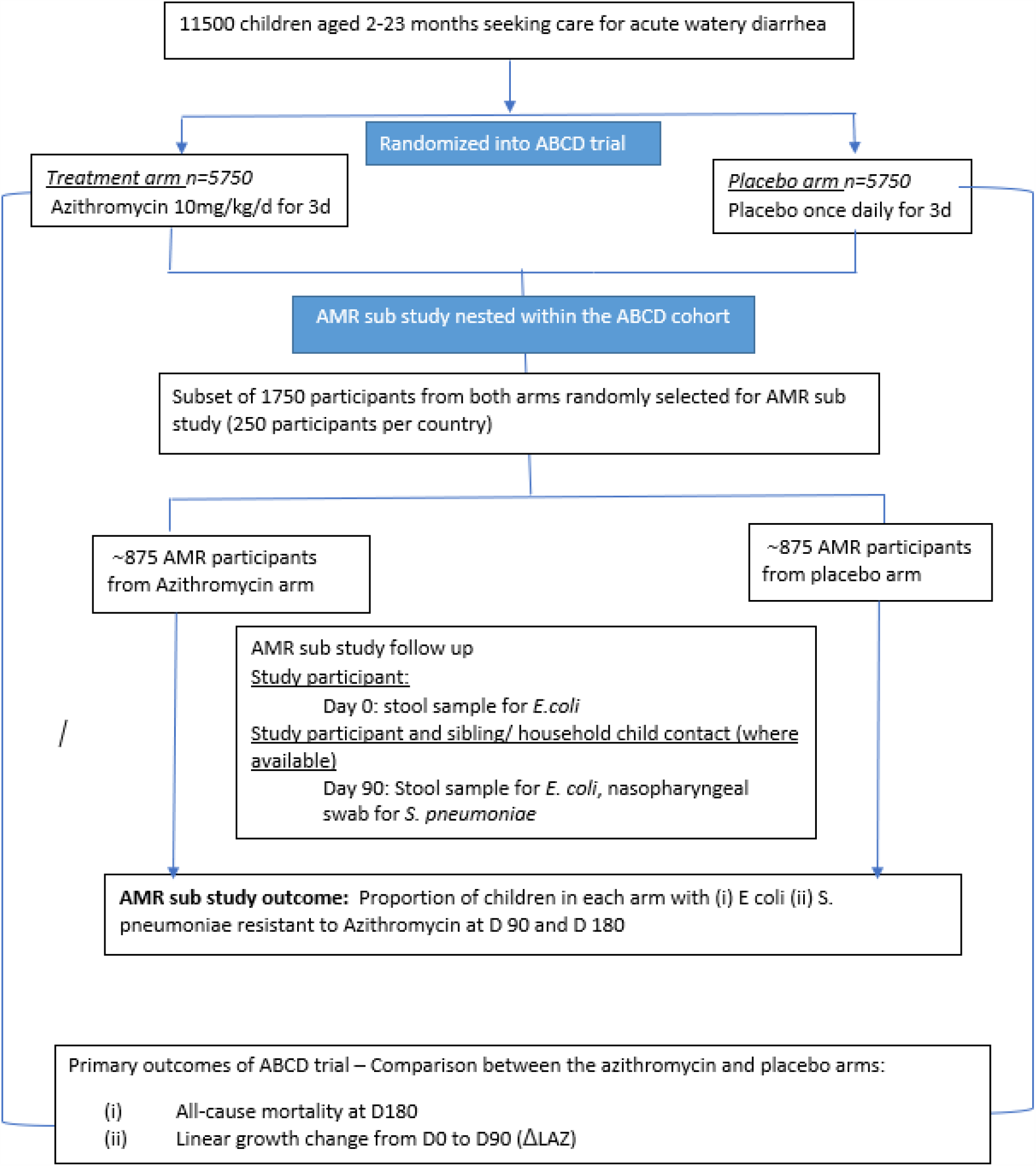
Antimicrobial resistance study nested within the ABCD trial

#### Study setting

The study will be conducted in health facilities in South Asia (Bangladesh, India and Pakistan) and sub-Saharan Africa (Kenya, Malawi, Mali and Tanzania). Within each country, 2-10 individual health facilities will be the sites of patient enrolment.

### Study procedures

#### ABCD Parent Study

The main ABCD trial protocol (not including the AMR sub study) is fully presented elsewhere^23^. In brief, children aged 2-23 months presenting to health facilities with diarrhea will be screened for enrolment. We enrolled young children at high risk of mortality in the 180 days following an acute diarrhea episode i.e. children with severe stunting, moderate malnutrition and/or dehydration. Children presenting with a history of bloody stool (as per caregiver report or health care worker observation) and children with another indication for antibiotic use (children presenting with presumed cholera (symptoms consistent with cholera and child presenting in a location with a declared cholera outbreak)), children with severe acute malnutrition (SAM), and/or children with signs of other infections requiring antibiotic treatment) were excluded. Participants receive the first dose of azithromycin or placebo in the health facility and a community health care worker directly observes the subsequent two doses on day 2 and day 3. All children are followed up at day 90 and day 180 to ascertain vital status, hospitalizations, his/her nutritional status (weight, length and Middle upper arm circumference MUAC). The primary outcomes of the trial are all-cause mortality during the 180 days post-enrolment and change in linear growth 90 days post-enrolment.

### AMR Sub Study

At ABCD study enrolment, 1750 randomly selected index children were recruited into the AMR sub-study. These participants were selected on the basis of systematic random sampling, so that every third child in the random ABCD sample was selected, during the first year of recruitment into the ABCD trial (2018). Samples (stool) were collected from the participating children at baseline (day 0), and from participating children and their siblings or close household contacts at day 90 and day 180. NP swabs were collected from participating children and their contacts at day 90 and day 180. Eligible contacts were children aged less than 60 months, who slept in the same household as the enrolled child for 5 of the last 7-nights, and who shared the same primary caregiver as the index child. If there are multiple contacts in the household who meet the eligibility criteria, then the preference for inclusion as a contact was given to the child closest in age and living in the same dwelling as the index child. Contacts were followed longitudinally through day 90 and day 180 to determine within-individual changes in resistance.

### Sample Collection and Processing

Stool samples will be collected from all participants enrolled in the AMR study at enrolment (index children only), day 90 and day 180 (index children and child contacts). If whole stool collection was unsuccessful, flocked rectal swabs (2) were collected. The stool sample or rectal swab is immediately placed in Cary-Blair transport media and transported to microbiology facilities (at temperatures between 2-8°C) within 24 hours of collection. NP swabs were collected from all participants enrolled in the AMR sub-study and from their contacts at day 90 and day 180 (but not at enrolment to reduce discomfort from the swabbing process at study entry). These are immediately be transferred into a 2ml vial containing Skimmed Milk Tryptone Glucose Glycerol (STGG)^24^. Vials will either processed or frozen at -80°C within 4 hours of collection.

### *E. coli* recovery protocol

Faecal samples stored in Cary Blair will be brought to room temperature and directly inoculated onto CHROMagar *E*.*coli* media (Becton Dickenson, USA). These will be checked for growth of *E. coli* after 24 hours of incubation at 35°C ±2°C. Suspected *E. coli* colonies, will be identified based on colony morphology and blue colouration, a single colony will be Gram stained. If the colony stains as a Gram-negative rod, a further suspected E. *coli* colony of the same morphology and colour will be transferred to MacConkey plates to obtain a pure culture. If, after 24 hours of incubation at 35°C ±2°C, non-lactose fermenting growth is shown, (usually pink or red growth), then the colonies will be sub-cultured onto a TSA (Tryptic soy agar) plate. A spot indole test will be carried out to confirm presence of *E. Coli* in the culture. Confirmed colonies of pure *E. coli* will be transferred from TSA plates into three 2 ml tubes containing Tryptic Soya Broth with 15% glycerol or Microbank storage beads. The vials will then be stored at -80°C, for future use and isolate recovery for Minimum inhibitory concentration (MIC) analysis.

### Nasopharyngeal Sample Processing and S. *pneumoniae* (SPN) recovery

Two hundred microliters of the inoculated STGG will be transferred into Todd Hewitt enrichment broth (5ml) containing 0.5 % yeast extract, supplemented with 1ml rabbit or sheep serum. This will be used to inoculate sheep blood agar plates supplemented with 0.5% gentamycin (SBG). After incubation for 18-24 hours at 35°C ±2°C in a CO_2_-incubator, the SBA will be examined for typical pneumococcal colonies; draughtsman shaped colonies surrounded by a zone of alpha-hemolysis. Growth may also occur in the form of large mucoid pneumococcal colonies for specific serotypes. Suspected pneumococcal colonies will be streaked onto SBG in confluent lines. After streaking, an optochin disc (5 μg optochin) will be placed in the streaked area and incubated in a CO_2_-incubator or candle-jar at 35°C ±2°C for 18-24 hours. Susceptibility to optochin (zone diameter >14 mm), would be confirmed as *S. pneumoniae*. Isolates identified as optochin resistant with zone diameter less than <14 mm, will be confirmed via bile solubility test. Pure cultures will be MIC tested either immediately or stored in tryptic soya broth with 15% glycerol at -80°C, for future MIC analysis.

### Methodology for MIC testing

#### E *coli*

Antimicrobial susceptibility testing (AST) for *E. coli* will be performed on the Beckman Coulter MicroScan, autoSCAN-4. This test is a minimized version of the broth dilution susceptibility test. Antimicrobial agents will be diluted in the required buffer to concentrations bridging the range of clinical interest. After inoculation of the broth with a standardized suspension of the organism at 35°C ±2°C for a minimum of 16-20 hours, the minimum inhibitory concentration (MIC) for *E. coli* will be determined by observing the lowest antimicrobial concentration showing inhibition of growth.

The standard bacterial culture will be scraped from the frozen pure colonies to MacConkey and TSA plates. The MacConkey plate will be used to rule out any contamination during storage or restoring of colonies. The plates will be incubated for 35°C ±2°C for 18-24 hours. Morphologically similar colonies will be picked with a sterile loop and emulsified in 3 ml of Microscan Inoculum Water (B1015-2) for a final turbidity equivalent to that of a 0.5 McFarland Turbidity Standard. 100 μL of the suspension obtained, is added to 25 mL of Inoculum Water with PLURONIC (Cat # B1015-7). Rehydration and inoculation will be performed using the RENOK system with inoculators-D. A final volume of 115 µL ± 10 µL will be used for a concentration between 3-7⨯10^5^ CFU/mL. Panels will be incubated for 16-20 hours to a maximum of 20 hours at 35°C ±2°C in an atmospheric non-CO_2_ incubator. Growth in the antimicrobial wells will be checked for turbidity. If turbidity is found in the negative control well or its absence is observed in the positive control well then experiment will be repeated. Panels will be processed and read in the Microscan auto SCAN4 instrument as per manufacturer instructions for all the listed antimicrobials (Appendix). Microscan instruments across all sites will be set to follow the CLSI 2020 guideline.

#### E-Test for *E.coli*

Because the Microscan Gram Negative panel does not include azithromycin, E-test strips will be used to obtain azithromycin MICs in *E.coli*. The E-test gradient strip will be used to directly quantify antimicrobial susceptibility in terms of discrete MIC values. Isolates recovered from frozen stocks will be suspended in 0.85% saline for a final turbidity equivalent to a 0.5 McFarland Turbidity Standard. Muller Hinton Agar will be smeared with the inoculum and the E-test strip placed on it for incubation in 35±2^°^C in a non-CO_2_ incubator for 16-20 hours. The MIC value will be read from the scale in terms of μg/mL where the pointed end of the ellipse intersects the strip.

#### S. pneumoniae

Pneumococcal isolates from the glycerol stock will be plated onto a TSA plate supplemented with 5% sheep blood. Plates will be incubated for 16-20 hours at 35°C ±2 °C at 5-10% Co2. Morphologically similar well isolated colonies will be emulsified in 3 ml of MicroScan inoculum water to create an inoculum equivalent to that of a 0.5 McFarland Turbidity Standard. 100 µl of the standardized suspension will be added to 25mLof Mueller Hinton broth with 3% lysed Horse blood broth. Rehydration and inoculation will be performed using a RENOK system inoculator-D. After inoculation the plates will be incubated for 20-24 hours at 35°C ±2°C in a non-CO_2_ incubator. After incubation, the plates will be checked for growth in the G wells. If turbidity is found in the negative or its absence is observed in the positive control well then experiment will be repeated. Visual readings will be reported on the LabPro software on a visual image of the MicroStrep plus panel. Detail of all the antimicrobials tested (including azithromycin) are presented in Appendix.

Patients and public were not involved in the design on the antimicrobial substudy.

### Data Analysis

#### Outcome definitions

Clinical Minimum inhibitory concentration (MIC) cut-offs corresponding to non-susceptibility from the 2020 Clinical and Laboratory Standards Institute (CLSI) will be used for defining azithromycin (and other antibiotic) resistance in isolates of *E.coli* and *S. pneumoniae*. (For azithromycin and E.coli, we will use the CLSI 2020 MIC break points for Enterobacteriales, as defined for S.enterica ser typhimurium, as CLSI 2020 does not publish an E.coli specific MIC break point). MICs exceeding the established cut-points corresponding to resistance and intermediate will be considered as “resistant” to maximize likelihood of detected potential signals of declining susceptibility. In the primary analysis, the denominator of these prevalence estimates will be all children enrolled in the AMR sub-study whose stool was submitted for AMR testing (irrespective of whether the bacteria were isolated). For the secondary analysis, we will limit comparisons to only those children in whom the bacterium was isolated.

#### Statistical Analysis and Sample Size

*<u>Objective</u> <u>1</u>*: To compare prevalence of azithromycin resistant *E.coli* and azithromycin resistant *S. pneumoniae* respectively in stool and NP swabs submitted for antimicrobial resistance testing at D90 and D180 among a random subsample of enrolled participants, by trial allocation arm. We then will test whether the upper limit of a one-sided 95% CI of the difference in prevalence of resistance at day 90 and day 180 in children assigned to the azithromycin and placebo group exceeds the non-inferiority margin of 10%. This margin has been chosen based on clinical acceptability among infectious disease experts.

In primary analyses, the prevalence denominator will be all children enrolled in the AMR sub-study, across all sites, whose stool for *E.coli* or NP swab for *S. pneumoniae* was submitted for antimicrobial sensitivity testing (irrespective of whether the bacteria were isolated). The non-inferiority hypothesis will be tested first using the continuity corrected χ2 test as suggested by Dunnett and Gent, Biometrics 1977^25^ for (i) the proportion of stools with azithromycin resistant *E. coli* at Day 90; (ii) proportion of stools with azithromycin resistant *E. coli* at Day 180; (iii) proportion of NP swabs with azithromycin resistant *S. pneumoniae* at Day 90; (iv) proportion of NP swabs with azithromycin resistant *S. pneumoniae* at Day 180. Children who were enrolled in the AMR study, who provided a sample, but whose sample could not be subject to an antibiotic sensitivity test because of external circumstances (COVID-19 related lockdowns of the lab), will be excluded from the denominator. If for any reason, sites are unable to complete testing of all their samples, random distribution of selection of specimens for testing will be difficult to confirm, though examination for systematic bias (e.g. by date range) will be undertaken. If bias is suggested in sites that have not completed testing, these sites will be excluded from hypothesis testing. But other remaining sites will be included in non-inferiority test as outlined above. In a secondary analysis, we will also explore the effect of azithromycin among the subgroup of children enrolled from African sites and Asian sites.

We will also examine the effect of azithromycin on isolates wherein the prevalence denominator will be defined as those children in whom the respective bacteria were isolated and tested (i.e. azithromycin resistance in E. coli as a proportion of E. coli isolates, and likewise for *S. pneumoniae*). Sites that have not completed testing as outlined above, will be dealt with as outlined above. For both the primary and secondary analyses, estimates of AMR prevalence will be adjusted for site and other covariates that are imbalanced between the two intervention groups of the AMR sub study.

We will also explore the change in resistance over time within individual participating children. This will be estimated using a binomial mixed effects model, as MICS will be reported categorically as resistant/sensitive. We will additionally report resistance patterns to other commonly used first and second-line antibiotics between the randomization arms in separate analyses.

*<u>Objective</u> <u>2</u>*: To compare prevalence of azithromycin resistant *E. coli* and azithromycin resistant *S. pneumoniae* respectively in stool and NP swabs submitted for antimicrobial resistance testing at Day90 and Day180 among the siblings or close household contacts (children under five years of age living in the same household under the care of the same primary caregiver) of a random sub-sample of enrolled participants, by trial allocation arm.

We will conduct the similar primary, secondary and exploratory analyses described for Objective 1 in the population of sibling or close household contacts (objective 2) with the same non-inferiority margin of 10%. We will only formally test for non-inferiority in the primary analyses of the prevalence

### Sample size estimation for AMR sub-study

The primary analysis of prevalence of azithromycin resistant *E. coli* in stool and *S. pneumoniae* in NP swabs will be done in all participants whose samples were submitted for antimicrobial resistance testing at Day 90 and Day 180.

However, due potential differences in resistance by region (Africa and Asia) we powered the study adequately assess non-inferiority by region. Therefore, the minimum number of participants required per treatment group in each region was calculated assuming an alpha of 0.05, 90% power, a non-inferiority margin of 10%, and a 25% prevalence of azithromycin resistance. This was based on baseline azithromycin resistance estimates in *E.coli* cultured from young children in Tanzania (16%) prior to mass drug administration of azithromycin^26^, in Bangladesh azithromycin resistance in *E.coli* in cases with diarrhea was estimated to be 27% ^27^. We also assumed 10% will be lost to follow-up and 10% will refuse to have the sample taken, leading to a required sample size for each region of 775 randomly selected children per region, of which 645 are assumed to provide samples for analysis. For the three sites in Asia, this translated to approximately 250 children randomly selected for the AMR sub-study per site. We decided to include the same number of children into the AMR sub study from each of the 4 African sites. The sample size for the primary objective (difference in AMR between the Azithromycin and placebo arm across all sites) and non-inferiority testing would therefore comprise the 1750 children recruited to the AMR study from each region,750 from the Asian sites and 1000 from the African sites. Therefore, in the primary analyses among all children, we expect a total of 1456 stools and NP swabs samples at each time point and we will >99% power to assess a non-inferiority margin of 10%.

In addition, we will also assess azithromycin resistance prevalence among siblings or close household contacts of ABCD participants at Day 90. Of the 1750 ABCD child participants randomly selected for the AMR sub-study, we assumed that 70% will have a potentially eligible sibling or close household contact and assumed 20% of these will refuse the sample collection. As a result, we assume 980 siblings or household contacts under 5 years. This will allow 99% power to assess a non-inferiority margin of 10% in the prevalence of resistant *E. coli* in stool and *S. pneumoniae* in NP swabs, among the participant siblings/contacts. In a secondary analysis, with 420 siblings or close household contacts providing samples at the Asian sites we will have >80% power to assess a difference of more than 10% between the treatment arms (non-inferiority margin). We will have ∼90% power for the same analysis with 560 siblings or close household contacts providing samples for the African sites.

### Conclusion

This study will provide valuable data informing policy makers on the cost, in terms of antibiotic resistance, of expanding azithromycin use for diarrhoea. This study will provide comparative data from a broad range of geographic and epidemiologic settings in South Asia and sub-Saharan Africa and will provide data describing temporal changes in resistance patterns over time. Additionally, as azithromycin is being considered for mass drug administration in high mortality settings, this study will also provide important individual-level data on the short and long-term consequences of azithromycin use in young children.

## Data Availability

This is a methods protocol paper, and therefore does not generate any data

## Acknowledgements

The ABCD team would like to thank the children and families that have participated in the trial to date. All the staff in all the participating sites are acknowledged for their dedication. The data management team at RTI international are acknowledged for supporting the data management function.

## Author contributions

All authors contributed to the drafting of the manuscript. All authors reviewed the manuscript for intellectual content and approved the final version of the report. No authors report conflicts of interest.

## Ethical approval

The trial has been approved by the WHO Ethics Review Committee and by the Ethics Committees of all participating sites in the seven countries – Bangladesh (Ethical Review Committee of icddr,b), India (Institutional Ethics Committee, Subharti Medical College & Hospital, Swami Vivekanand Subharti University), Kenya (Kenya Medical Research Institute Ethical Review Committee and the University of Washington Institutional Review Board), Malawi (University of Malawi College of Medicine Research Ethics Committee), Mali (Comite d’Ethique de la FMPOS (ERC at USTTB, University of Maryland, Baltimore Research Ethics Committee), Pakistan (Aga Khan University, Ethical Review Committee), Tanzania (Muhimbili University of Health and Allied Sciences Senate Research and Publications Committee; Tanzanian Food and Drug Administration; National Institute for Medical Research), Boston (Boston Children’s Hospital Institutional Review Board). Each updated version of the approved protocol will be submitted to the ethical committees above.

Written informed consent (parents/caregivers) is obtained by study staff before any trial procedures are carried out and participants’ confidentiality is maintained throughout the trial in line with the standard ICH-GCP principles.

## Appendix 1

| Antimicrobial agent | E.coli* | S. pneumoniae# |
| --- | --- | --- |
| Amikacin | X |  |
| Amoxicillin / K Clavulanate | X | X |
| Ampicillin | X | X |
| Ampicillin /Sulbactam | X |  |
| Azithromycin |  | X |
| Aztreonam | X |  |
| Cefepime | X | X |
| Cefotaxime | X | X |
| ESβL confirmation-Cefotaxime / K Clavulanate | X |  |
| Cefoxitin | X |  |
| Ceftazidime | X |  |
| ESβL confirmation-Ceftazidime / K Clavulanate | X |  |
| Cefuroxime | X | X |
| Cephalothin | X |  |
| Chloramphenicol | X | X |
| Ciprofloxacin | X |  |
| Clarithromycin |  | X |
| Clindamycin |  | X |
| Colistin | X |  |
| Daptomycin |  | X |
| Doripenem | X |  |
| Ertapenem | X |  |
| Erythromycin |  | X |
| Fosfomycin | X |  |
| Gentamicin | X |  |
| Imipenem | X |  |
| Levofloxacin | X | X |
| Linezolid |  | X |
| Meropenem | X | X |
| Minocycline | X | X |
| Moxifloxacin |  | X |
| Nalidixic Acid | X |  |
| Nitrofurantoin | X |  |
| Norfloxacin | X |  |
| Penicillin |  | X |
| Piperacillin | X |  |
| Piperacillin / Tazobactam | X |  |
| Pristinamycin |  | X |
| Rifampin |  | X |
| Tetracycline | X | X |
| Tigecycline | X |  |
| Tobramycin | X |  |
| Trimethoprim / Sulfamethoxazole | X | X |
| Vancomycin |  | X |

\* MicroScan dried Gram Negative MIC panel 44 - NM 44 B/20 (Cat # B1016-175)

